# Screening tools used in primary health care settings to identify health behaviours in children (birth – 16 years); A systematic review of their effectiveness, feasibility and acceptability

**DOI:** 10.1101/2023.03.02.23286714

**Authors:** Dimity Dutch, Lucinda Bell, Dorota Zarnowiecki, Brittany J Johnson, Elizabeth Denney-Wilson, Rebecca Byrne, Heilok Cheng, Chris Rossiter, Alexandra Manson, Eve House, Kamila Davidson, Rebecca K Golley

## Abstract

**Background:** Child health behaviour screening tools used in primary health care have potential as a transformative and effective strategy to support growth monitoring and the early identification of suboptimal behaviours to target strategies for intervention. This systematic review aimed to examine the effectiveness, acceptability and feasibility of child health behaviour screening tools used in primary health care settings.

**Methods:** A systematic review of studies published in English in five databases (CINAHL, Medline, Scopus, PsycINFO and Web of Science) prior to July 2022 was undertaken using a PROSPERO protocol and PRISMA guidelines. Eligible studies: 1) described screening tools for health behaviours (dietary, physical activity, sedentary or sleep-related behaviours) used in primary health care settings in children birth to 16 years of age; 2) reported their acceptability, feasibility or effectiveness on child or practitioner behaviour or 3) reported implementation of the screening tool. Study selection and data extraction were conducted in duplicate. Results were narratively synthesised.

**Results:** Of the 7145 papers identified, 22 studies reporting on 14 unique screening tools were included. Four screening tools measured diet, physical activity, sedentary and sleep behaviours domains, with most screening tools only measuring two or three behaviour domains. Ten studies reported screening tools were effective in changing practitioner self-reported behaviour, knowledge, self-efficacy and provision of health behaviour education. Administration of screening tools varied across studies including mode, timing and caregiver or practitioner completion. Implementation strategies described included practitioner training and integration into electronic medical records. Practitioners and caregivers identified numerous benefits and challenges to screening; however, child views were not captured.

**Conclusions:** Few screening tools exist to facilitate comprehensive screening of children’s health behaviours in primary health care. This review highlights the potential of health behaviour screening as an acceptable and feasible strategy to comprehensively assess and provide early intervention for children’s health behaviours in primary health care settings.

**Potential conflicts of interest:** All authors have no conflicts of interest to declare.

## 1. INTRODUCTION

Poor diet quality, inadequate physical activity, and poor sleep habits are key modifiable health behaviours contributing to significant health and economic burden globally. Over one third (38%) of total chronic disease burden is potentially avoidable due to modifiable risk factors^1, 2^. Health behaviours are established during childhood and adolescence and can have a significant influence on health across the life course ^3, 4, 5^.Therefore, identification of poor health behaviours and intervention in early life is critical to support lifelong health ^6, 7^.

Primary Health Care is defined by the World Health Organisation (WHO) and the United Nations Children’s Fund (UNICEF) as being “*“a whole-of-society approach to health that aims at ensuring the highest possible level of health and well-being and their equitable distribution by focusing on people’s needs and as early as possible along the continuum from health promotion and disease prevention to treatment, rehabilitation and palliative care, and as close as feasible to people’s everyday environment”*^*8*^ and is often the first point of contact to the health care system for families of young children. PHC is therefore an opportunistic and important setting for promotion of, and early intervention for positive health behaviours in childhood and adolescence. Primary Health Care is a trusted, valued and accessible setting for children and their families, with key responsibilities in screening for disease risk factors and providing counselling for families^9, 10, 11^. Current recommended practice within PHC is to identify children with or at risk of overweight or obesity, as a proxy for poor health behaviours, based on growth monitoring, with or without brief advice for health behaviours^12, 13, 14, 15^. However, several international systematic reviews have found a lack of high-level evidence to support the effectiveness of routine growth monitoring as a screening tool in practice, and its benefit on child health^16, 17, 18^. Further, practitioners have difficulty plotting and interpreting growth charts to inform practice, resulting in potentially inappropriate or ill-informed advice^19^ while caregivers are often not receptive to weight-focussed conversations^20, 21, 22^. As a result, there is opportunity to utilise measures of diet quality, physical activity, sedentary behaviours and sleep habits to better understand a child’s unique health behaviours and provide tailored advice to families.

‘Gold standard’ methods of measuring health behaviours such as accelerometry and diet histories can be time consuming and are therefore not feasible in time-poor settings such as PHC^23, 24^. Brief screening tools can be a time-efficient and cost-effective method of assessing health behaviours, allowing for identification of specific target behaviours to inform individualised counselling and intervention. Incorporation of screening for health behaviours into PHC practice addresses limitations experienced with current growth monitoring practice and provides greater insight into child health, beyond weight status. The interrelated nature of health behaviours means it is important to identify and manage behaviours as they exist collectively, rather than in isolation^25, 26, 27^. Thus, brief screening tools that measure diet, activity, sedentary and sleep-related behaviours pose an effective strategy to support long-term population health and a more cost-effective and sustainable PHC system.

To enable screening of children’s health behaviors in PHC, valid and reliable screening tools are needed. A systematic review by Byrne and colleagues identified and described the validity and reliability of 12 brief screening tools to measure health behaviours in children in the first 5 years of life^28^. However, none of the included screening tools measured all four health behaviour domains (dietary intake, physical activity, sedentary behaviour, and sleep), and few were used or evaluated in PHC settings. Thus, their suitability for application in this setting is unknown. Further tools were identified in a recent systematic review by Krijger and colleagues, which described 41 unique screening tools to measure lifestyle behaviours in children aged 0-18 years in community settings^29^. However, the tools described in this review ranged in length, with several tools >25 items in length, impacting their suitability for use in the time-poor PHC setting. Additionally, these reviews did not address: post-screening actions (i.e., counselling or referral pathways) essential for enabling positive behaviour change; caregiver or practitioner acceptability and feasibility; or the effectiveness of child health behaviour screening in PHC settings, which is required to understand if brief screening is suitable for widespread adoption. A gap also exists in knowledge regarding the implementation strategies, and the tools and resources required to embed health behaviour screening into routine PHC practice.

Thus, the aim of this systematic review was to identify and describe screening tools used in PHC settings that measure health behaviours in children from birth to 16 years, and to determine their effectiveness in identifying health behaviours and changing practitioner knowledge, attitudes and/or practice. The secondary aims were to understand practitioners’, caregivers’ and children’s views of health behaviour screening tools, and the training and resources required to support implementation of health behaviour screening within practice.

## 2. METHODS

This systematic review followed a prospectively prepared protocol (PROSPERO International prospective register of systematic reviews: registration number: CRD42022340339 https://www.crd.york.ac.uk/prospero/) and is reported using the Preferred Reporting Items for Systematic Reviews and Meta-Analyses (PRISMA) guidelines for systematic reviews^30^.

### 2.1 Search strategy and information sources

A comprehensive and systematic search of five electronic databases (CINAHL, Medline, Scopus, PsycINFO, Web of Science) was undertaken in July 2022 to identify screening tools used with children and/or caregivers in the PHC setting for the identification of health behaviors (i.e., diet, physical activity, sedentary behaviour and sleep). Search terms were pilot tested, refined and tailored to each database in consultation with an academic librarian. Keywords and subject headings were organised into three categories, (i) population (e.g. infant, toddler, preschool, child, youth, adolescent, pediatric) AND (ii) context (e.g. primary health care, family practice, general practitioner, health professional) AND (iii) concept (e.g. screen/screener/screening, questionnaire, survey checklist, detect, identify, diagnosis, decision support systems, decision making). No publication date limits were applied. The full search strategy used in one database is presented in supplementary file 1.

### 2.3 Eligibility criteria

#### Types of studies

Included studies reported on empirical research, including randomized controlled trials, experimental studies, non-randomised comparison studies, pre-post designs, and qualitative research. Reviews, commentaries and letters to the editors, as well as dissertations and conference abstracts, were excluded.

#### Participants

Eligible participants included children aged ≤16 years of age and their caregivers, and PHC practitioners (e.g., practice managers, general practitioners, nurses). Studies that included children over 16 years of age were eligible provided the mean age was ≤16 years of age. This child age range was chosen as a child aged 16 years and older can consent to their own medical treatment^31^. For this review, caregiver is used to describe parents and other primary caregivers.

#### Concept

The concept of interest was screening tools (including decision support tools, diagnostic tools) for at least one child health behaviour or caregiving practices relating to diet, physical activity, sedentary behaviour, and sleep, such as rules and routines regarding family meals and screen use. Studies could examine the screening implementation approach, metrics of use, participant views including acceptability, attitudes or changes in practitioner screening behaviour. Screening tools could be delivered via any mode (e.g., paper or online) and be completed by any of the above participant groups (i.e., children, caregivers, practitioners). Studies were excluded if the screening tool focused solely on physical examination or diagnosis, assessed behavioural outcomes of weight loss interventions or the study used the screening tool to assess study eligibility only.

#### Context

Eligible studies were undertaken in any PHC setting internationally, including general practice, maternal and child health services, community health or Indigenous health services. Studies where the screening tool was used by specialists or services where children are referred for assessment or treatment of overweight were excluded.

### 2.3 Selection process

Study selection was undertaken using the web-based systematic review software Covidence^32^ by DD, HC, RB, CR, DZ, KD and AM. Studies were screened in duplicate against the a priori defined inclusion and exclusion criteria in two stages: (1) title and abstract screening, (2) full text screening of remaining articles. Any discrepancies were resolved by discussion. Reference lists of included articles and relevant reviews were also hand-searched to identify any additional relevant studies, which were subsequently checked for eligibility against the inclusion and exclusion.

### 2.4 Data extraction and risk of bias assessment

Data extraction was performed by one reviewer (DD) using a standardized review-specific data extraction table that had been piloted with selected studies prior and refinements made to ensure consistency in the extraction process across studies. Following data extraction of the first 10% of included papers by two reviewers (DD and Research Assistant), further amendments were made.

Data extracted included: author, year, study title; study details (study design, duration, setting) (Table 1); population characteristics (number of participants, child age, primary health care practitioner role, number of primary health care centres) (Table 1); screening tool characteristics (name, number of items, health behaviours addressed, administration method) (Table 2); changes in practitioner behaviour (Table 3); PHC practitioner views on screening tools (Figure 2); caregiver views on screening tools (Table 4); and practitioner-identified training and resource needs (Table 5). If the eligible screening tool was not available, corresponding authors were contacted via email to seek a copy for data extraction purposes.

**Table 1:** Summary of included studies.

| Study details | Intervention details | Child + Caregiver Population | PHC Practitioner Population | MMAT Score |
| --- | --- | --- | --- | --- |
| *First author (Year)* | *Study design* | *Child age<sup>a</sup>* | *Practitioner sample size* | *Out of 100%* |
| *Country* | *Intervention period/Study length* | *Child sample size* | *Number of PHC clinics* |  |
| Beno (2005)<sup>35</sup> | Intervention with follow up qualitative questionnaire and focus groups | Child age N/R | Practitioners n = 76<br>PHC Clinics n = 9 | 20% |
| United States | 6-months |  |  |  |
| Hinchman (2005)<sup>39</sup> | Delayed-control design | Children 5-18 years<br>Children n = 660 | Practitioners n=101<br>PHC Clinics n = 9 | 40% |
| United States | 6-months |  |  |  |
| Dunlop (2007)<sup>50</sup> | Medical Record Abstraction | Children 2-17 years<br>Children n = 1348 | Practitioners n = 38<br>PHC Clinics n = 6 | 80% |
| United States | 6-months |  |  |  |
| Woolford (2009)<sup>51</sup> | Mixed Methods | Children 2-5 years | Practitioners n = 15<br>PHC Clinics N/R | 20% |
| United States | 12-months |  |  |  |
| McKee (2010)<sup>49</sup> | Qualitative evaluation of pilot intervention | Children 22-59 months<br>Caregiver n = 18 | PHC Clinics = 3 | 60% |
| United States | Intervention period N/R |  |  |  |
| Watson-Jarvis (2011a)<sup>52</sup> | Descriptive cross-sectional survey | Child age N/R<br>Caregiver n = 412 | Practitioners n = 26<br>PHC Clinics n = 2 | 20% |
| Canada | 5-months |  |  |  |
| Watson-Jarvis (2011b)<sup>53</sup> | Descriptive cross-sectional survey | Children 3-≥6 years<br>Caregiver n = 438 | PHC Clinics n = 2 | 60% |
| Canada | 5-months |  |  |  |
| Andrade (2020)<sup>48</sup> | Mixed Methods | Children <17-72 months<br>Children n = 280 | Practitioners n = 5<br>PHC Clinics n = 5 | 40% |
| Canada | 12-months |  |  |  |
| Christison (2014)<sup>45</sup> | Prospective, non-randomized, observational study | Children 4-16 years<br>Children n = 100 | Practitioners n = 7<br>PHC Clinics n = 1 | 20% |
| United States | 14-weeks |  |  |  |
| Herbenick (2018)<sup>46</sup> | Evidence-based practice design | Children 4-11 years<br>Children n = 27 | PHC Clinics n = 1 | 20% |
| United States | 10-weeks |  |  |  |
| Bailey-Davis (2019)<sup>34</sup> | Quasi Experimental | Children 2-9 years<br>Children n = 10,647 | PHC Clinics n = 20 | 40% |
| United States | 12-months |  |  |  |
| Gance-Cleveland | Study design N/R | Child age N/R | Practitioners n = 14 | 20% |
| (2014)<sup>58</sup> | 8-months | Children n = 3,215 | PHC Clinics n=12 |  |
| United States |  |  |  |  |
| Park (2015)<sup>41</sup> | Uncontrolled pilot intervention study with questionnaire and semi-structured interviews | Children 5-18 years<br>Child mean age 10.7±2.6 years<br>Children n = 14<br>Caregiver n = 12 | Practitioners n = 4<br>PHC Clinics n = 4 | 20% |
| United Kingdom | 6-months |  |  |  |
| Sharpe (2016)<sup>43</sup> | Quality improvement study | Children 3-16 years<br>Children n = 41<br>Caregiver n = 41 | PHC Clinics n=1 | 20% |
| United States | 6-months |  |  |  |
| Polacsek (2009)<sup>42</sup> | Quasi experimental | Children 5-18 years<br>5-11years = 56%<br>12-17 years = 44%<br>Children n=600<br>Caregiver n=539 | Practitioners n=31<br>PHC Clinics n=19 | 20% |
| United States | 18-months |  |  |  |
| Gibson (2016)<sup>38</sup> | Retrospective and postintervention chart reviews | Preintervention child mean age<br>13.1±3.8 years<br>Children n = 134 | PHC Clinics n=2 | 60% |
| United States | 6-weeks |  |  |  |
| Camp (2017)<sup>37</sup> | Mixed Methods | Children 2-9 years<br>Children n = 601 | Practitioners n = 12<br>PHC Clinics n = 2 | 20% |
| United States | 8-weeks |  |  |  |
| Camp (2020)<sup>36</sup> | Mixed Methods | Children 2-9 years<br>Children n = 425 | Practitioners n = 12<br>PHC Clinics n = 2 | 20% |
| United States | 6-weeks |  |  |  |
| Karacabeyli (2020)<sup>40</sup> | Preintervention and postintervention observational mixed methods<br>9 months (Community A)<br>12 months (Community B) | Children age N/R | Practitioners n = 21<br>PHC Clinics n = 6 | 20% |
| Canada |  |  |  |  |
| Savage (2018)<sup>47</sup> | Protocol for a Randomised Controlled Trial | Children 0-6 months<br>Sample size aim:<br>n = 290 mother-infant dyads | PHC Clinics N/R | 20% |
| United States | 7-months |  |  |  |
| Shook (2018)<sup>44</sup> | Cross-sectional review of electronic medical records | Children 2-18 years<br>Children n = 24,255 | PHC Clinics n = 1 | 80% |
| United States | 3-years |  |  |  |
| Williams (2020)<sup>59</sup> | Mixed Methods | Children 3-17 years | Practitioners n = 44<br>PHC Clinics n = 2 | 20% |
| United States | 10-months |  |  |  |
| Abbreviations: MMAT: Mixed Methods Assessment Tool<sup>33</sup>, MMAT scored out of 100%, 20% per question, higher % score indicating higher quality study; N/R: Not reported |  |  |  |  |
| <sup>a</sup>Child age as reported in the study |  |  |  |  |

**Table 2:**
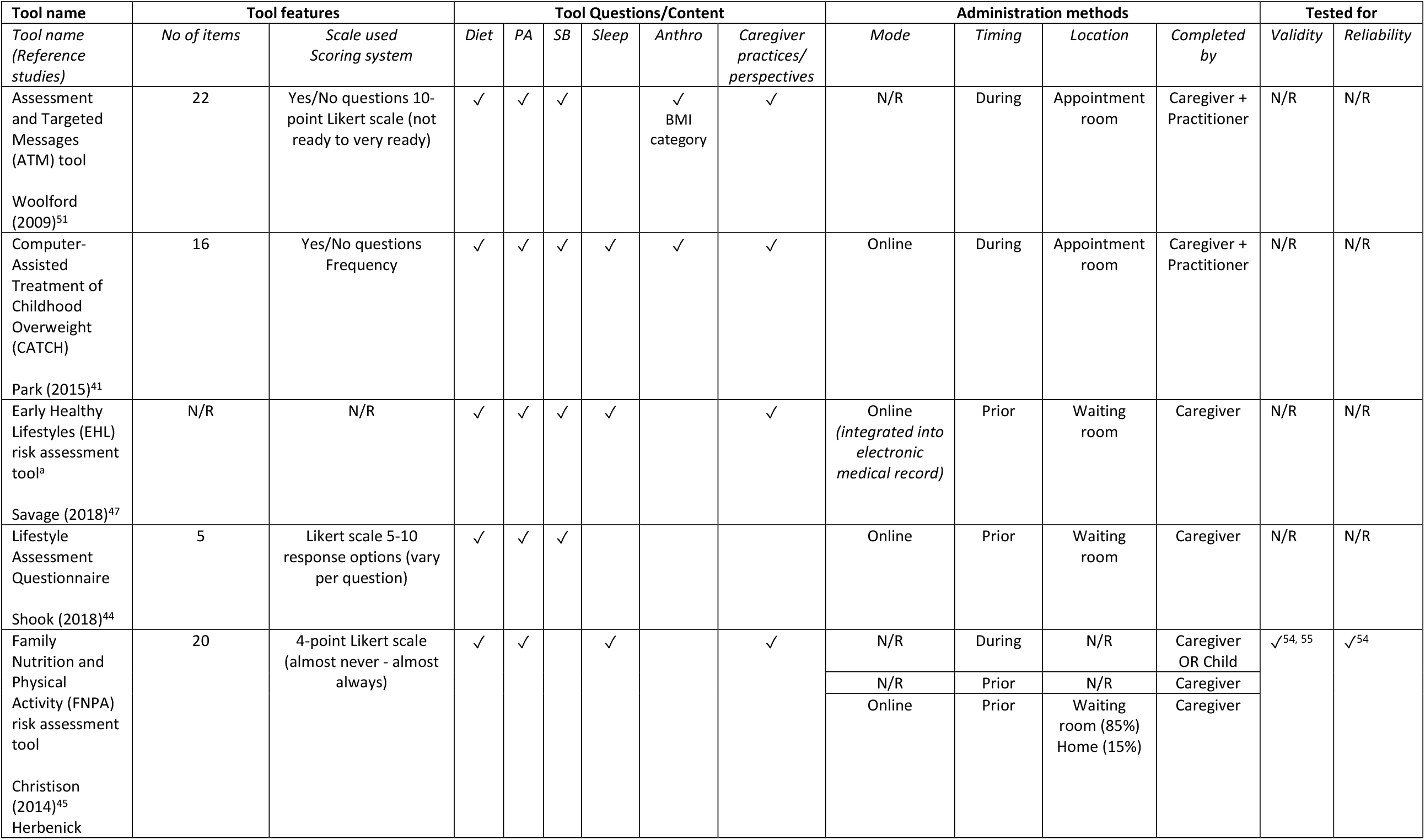
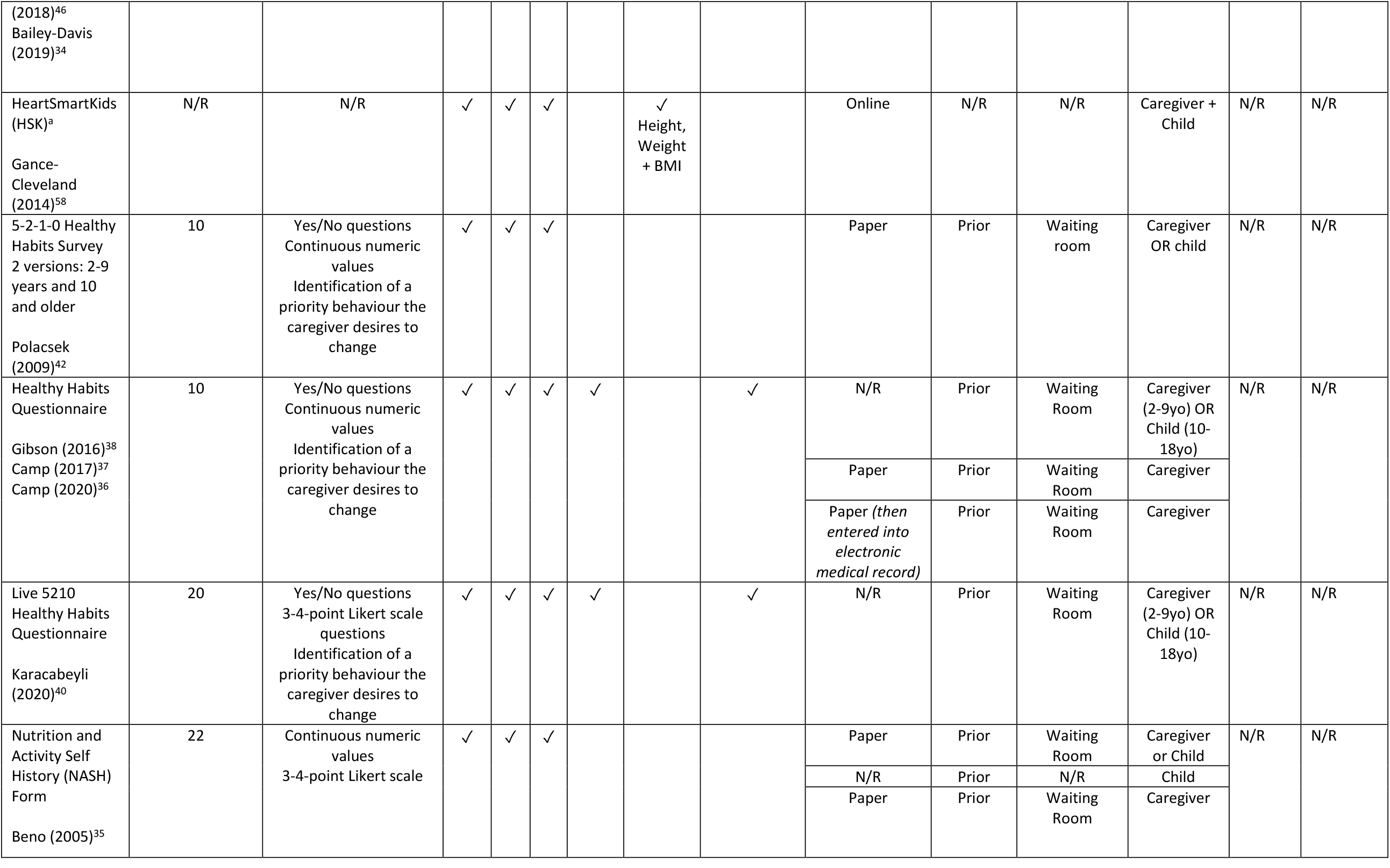
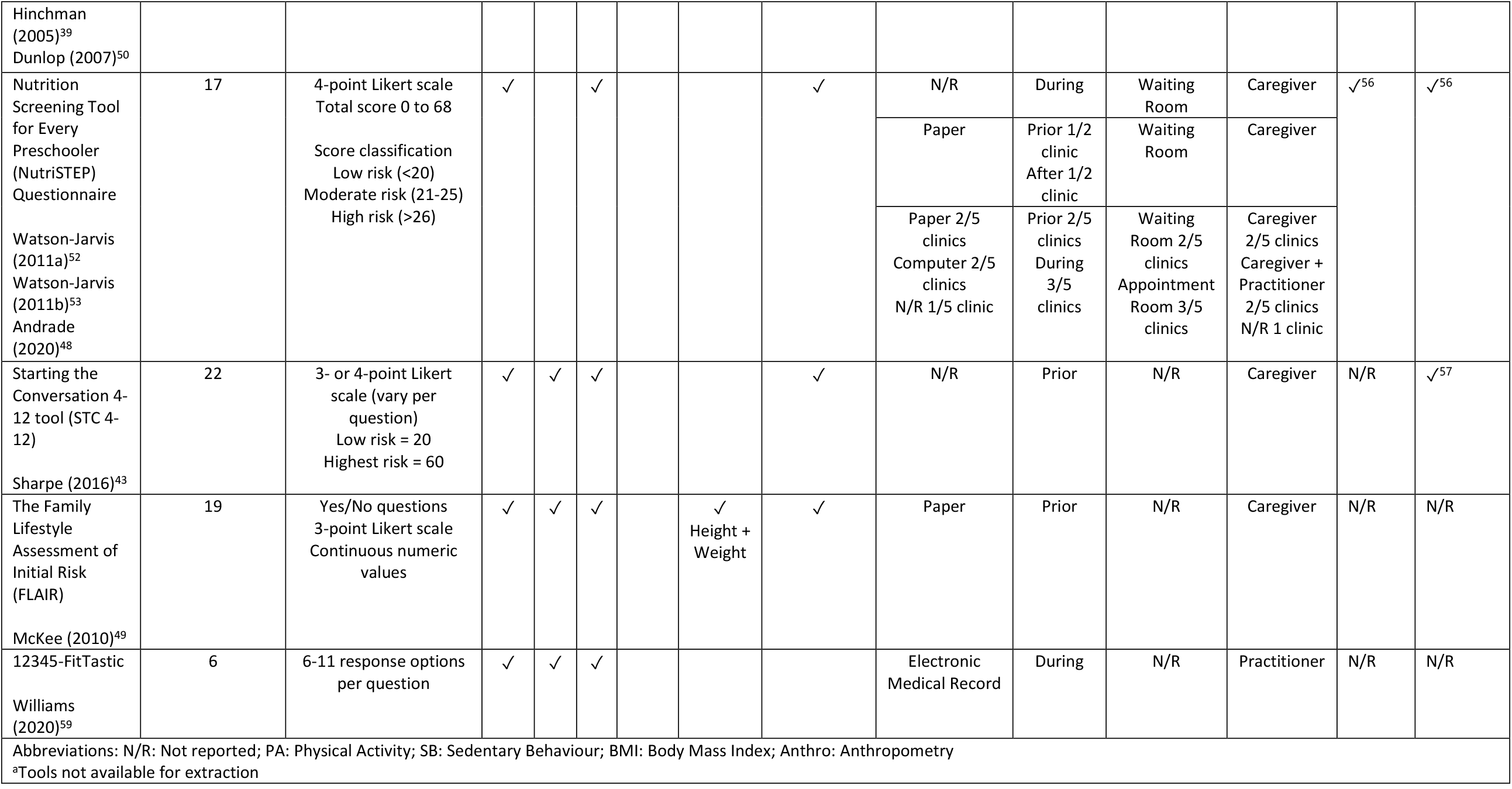
Characteristics of health behaviour screening tools identified for children in primary health care settings.

**Table 3:**
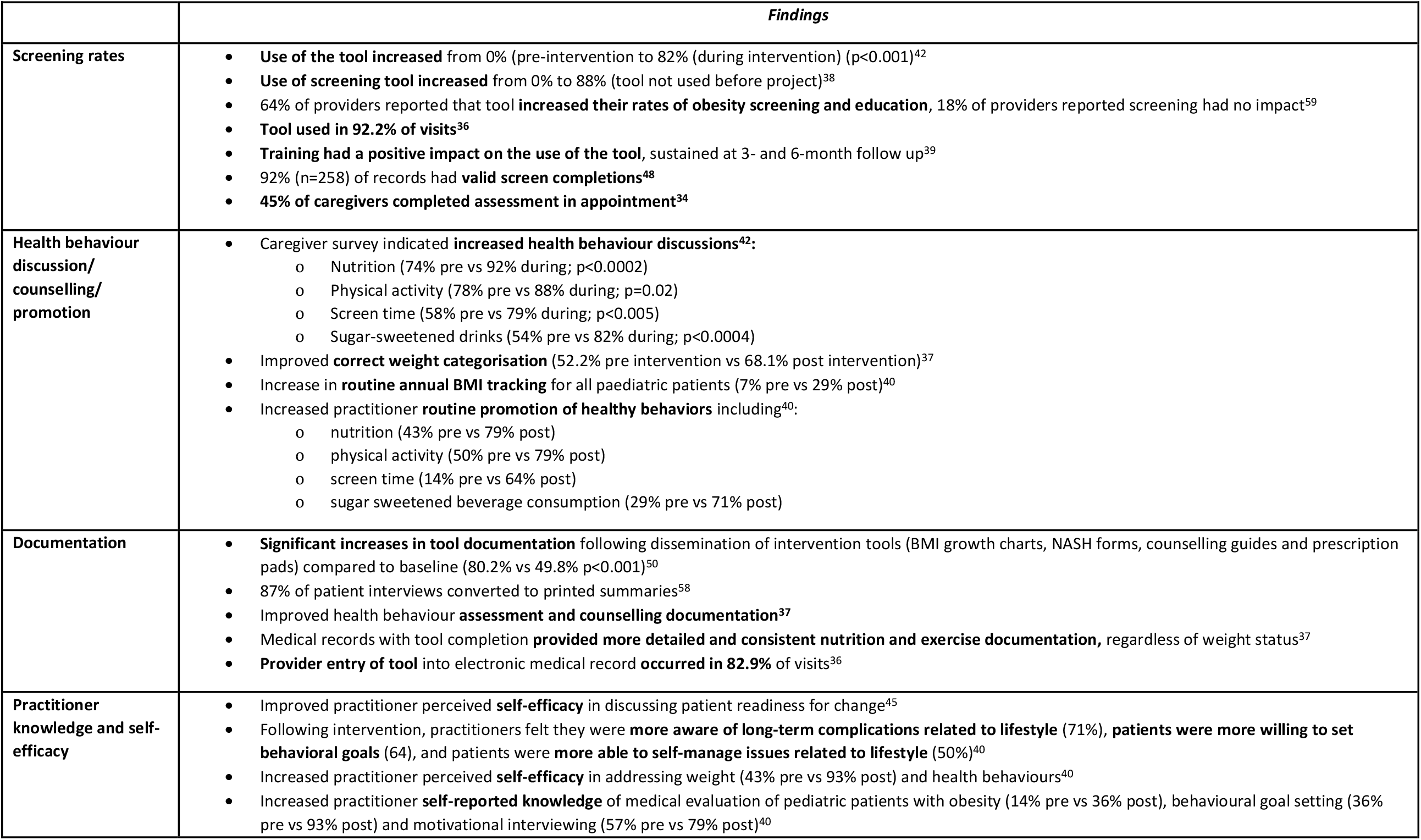
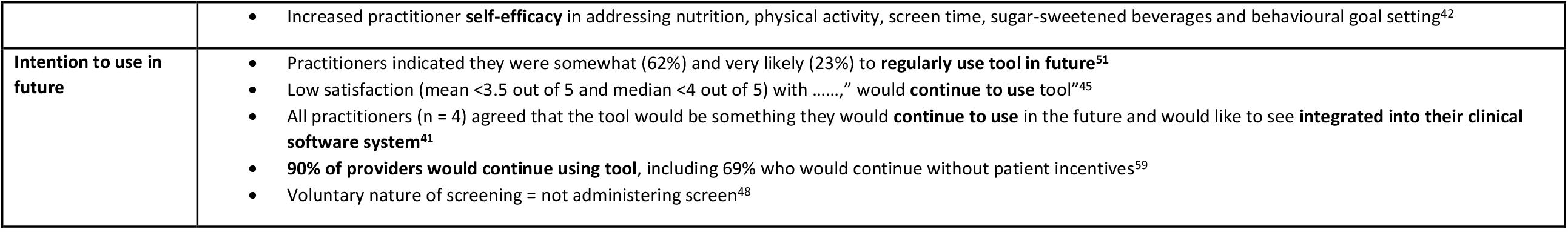
Changes in practitioner behaviour, knowledge and practice in health behaviour screening.

**Table 4:** Practitioner identified training and resources needs alongside health behaviour screening tool.

|  |  |
| --- | --- |
| **Training** | Training to providers about the tool<sup>48, 50</sup><br>Skill building training<sup>35</sup><br>Training to providers about how to prioritise and assess most significant behaviours<sup>58</sup><br>Affordable and practical in-service training<sup>39</sup><br>Training and technical assistance<sup>48</sup> |
| **Practitioner Resources** | More tangible support such as a structured program of activities + follow up consultations to monitor patients<sup>41</sup><br>Behaviour change list + Examples of exercise + healthy meal options for children<sup>51</sup><br>Key primer booklet<sup>48</sup><br>Access to ready-to-use resources alongside the screening tool<sup>40</sup><br>Decision support chart as part of resource toolkit<sup>38</sup> |
| **Electronic Medical Records** | Integration of tool into electronic medical records, automatic calculation of assessment<sup>41, 45</sup><br>Integration of reminders into EMRs<sup>48</sup> |
| **Dietitian support** | Onsite nutritionist/dietitian available for drop-in follow-up visits<sup>52</sup><br>Registered dietitian roles<sup>48</sup> |
| **Administrative support** | Administrative staff roles<sup>48</sup><br>Practitioners depended on administrative staff to administer the screening tool and implementation sustainability was contingent on capacity of front-end administrative staff<sup>40</sup> |
| **Patient education Resources** | Educational resources<sup>48</sup> |

**Figure 1.**
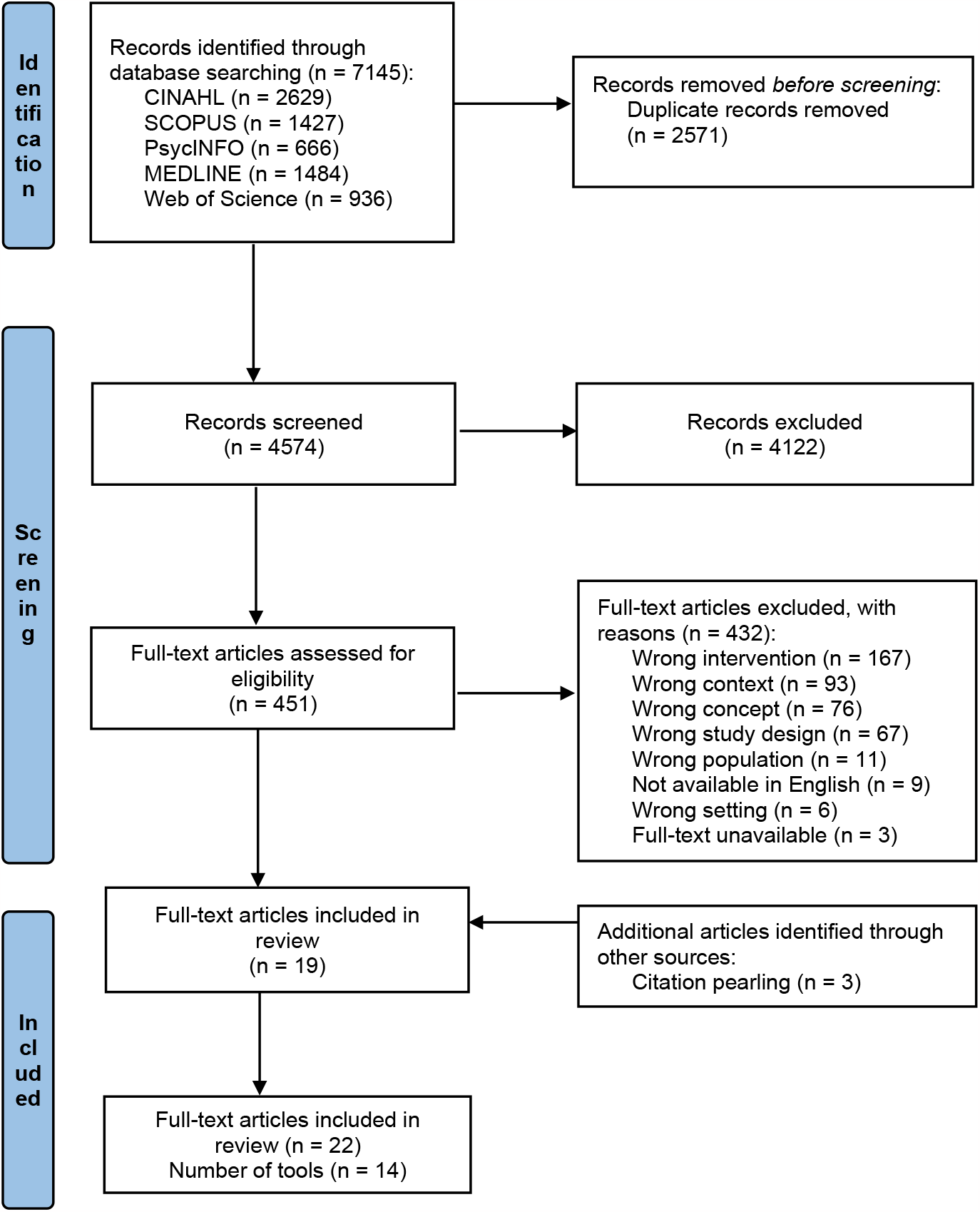
PRISMA statement flow diagram.

**Figure 2a:**
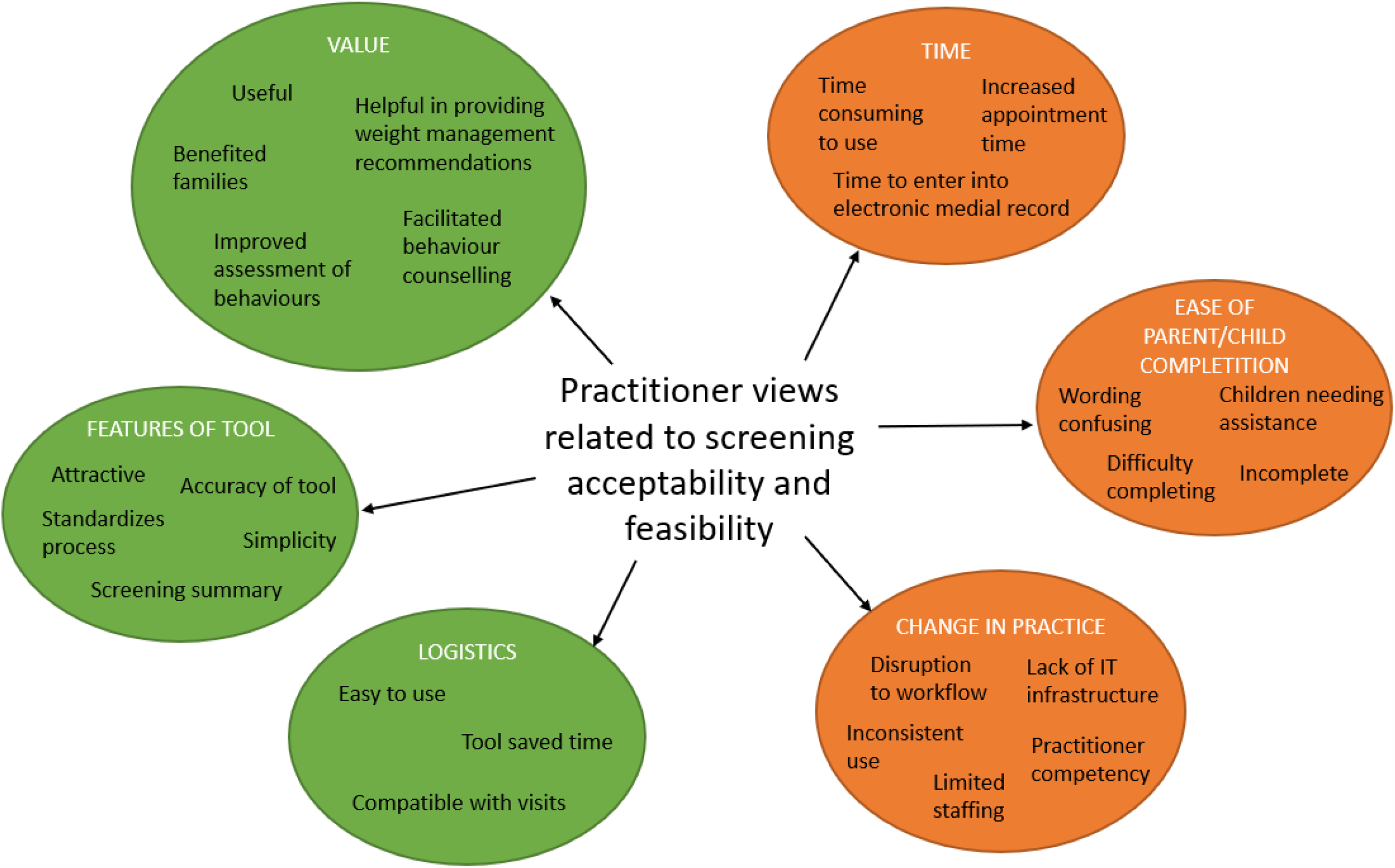
Practitioner views related to health behaviour screening acceptability and feasibility (n=14 studies)

**Figure 2b:**
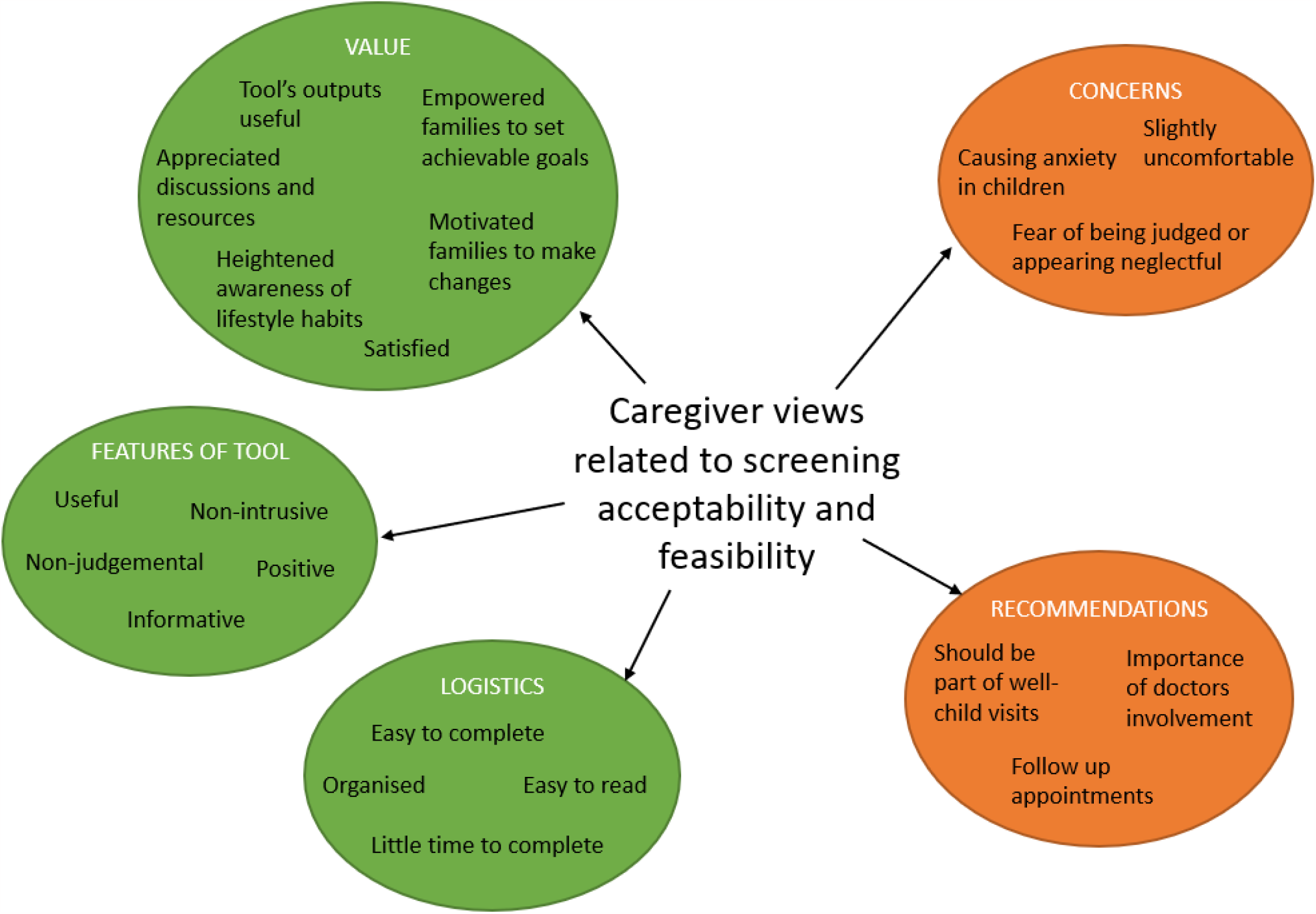
Caregiver views related to health behaviour screening acceptability and feasibility (n=8 studies)

Risk of bias assessment was undertaken with the Mixed Methods Appraisal Tool (MMAT)^33^ by two reviewers (DD and EH), which assesses study quality on five domains for five empirical study designs: (1) Qualitative, (2) Quantitative randomised controlled trials, (3) Quantitative non-randomised, (4) Quantitative descriptive, and (5) Mixed methods.

### 2.5 Data synthesis

A narrative synthesis approach was used in this review due to the range of different outcome measures reported in the included studies and due to the inclusion of both qualitative and mixed methods studies. The narrative synthesis of findings was structured to address the primary and secondary aims. Synthesis was organised into five key components: 1) description of available screening tools; 2) effectiveness of screening tools for changing PHC practitioner knowledge, attitudes and practice; 3) acceptability and feasibility of tools for a) PHC practitioners and b) caregivers and children; 4) training and resources required for implementation of screening tools.

## 3. RESULTS

### 3.1 Search results and characteristics of included studies

Database searching identified 7145 unique records of which 19 met the review criteria (Figure 1). An additional three eligible studies were identified through citation pearling. The final 22 studies included in this review were undertaken in the United States (US) (n = 17), Canada (n = 4) and the United Kingdom (UK) (n = 1) (Table 1). Studies were predominately non-controlled interventions or quality improvement projects^34, 35, 36, 37, 38, 39, 40, 41, 42, 43^, ranging in duration from 6 weeks^36, 38^ to 3 years^44^. The number of primary health care clinics included in a given study varied from one^43, 44, 45, 46^ to 20 clinics^34^. PHC practitioners included nurses, dietitians, physicians and paediatricians, as well as clinic staff, such as clerks and managers. Children included in the studies ranged in age from 0-6 months^47^ up to 18 years (e.g., 2-18 years), with only three studies including children aged <24 months^47, 48, 49^ and most studies including children >2 years of age (n= 17). Overall, MMAT scores were mixed, with 14 studies reporting low risk of bias in one of five domains, receiving a score of 20%. Only two studies^44, 50^ reported low risk of bias in four of five domains (score of 80%). None received a score of 100% (low risk of bias in all five domains) (Table 1 and Supplementary Table 1).

### 3.2 Characteristics of screening tools

Fourteen unique screening tools were identified across the 22 studies (Table 2). Four screening tools were not available in publication data – corresponding authors were contacted, of whom two responded to provide two screening tools as part of data extraction and synthesis: 5-2-1-0 Healthy Habits Survey^42^ and The Family Lifestyle Assessment of Initial Risk (FLAIR)^49^. Tools ranged in length from 5^44^ to 22 items^35, 39, 43, 50, 51^ and were completed by patients (caregiver, or caregiver and child), practitioners, or both, using various administration methods (paper, online or computer, electronic medical record-based), timing (during or, prior to, consultation), and locations (home, waiting room, appointment room). Four tools addressed all four health behaviours of diet, physical activity, sedentary behaviour and sleep: Computer-Assisted Treatment of CHildhood overweight (CATCH)^41^; Early Healthy Lifestyles (EHL)^47^; Healthy Habits Questionnaire (HHQ)^36, 37, 38^; Live 5-2-1-0 HHQ^40^; however, there was no evidence of these tools being tested for validity or reliability. Most tools (n = 9) addressed the three health behaviour domains of diet, physical activity and sedentary behaviour. One tool^48, 52, 53^ addressed only two health behaviour domains, diet and sedentary behaviour. Four tools addressed anthropometry (height, weight, BMI or BMI category). Nine tools measured caregiving practices or their perspectives related to their child’s health behaviours. Two tools have been tested for both validity and reliability^54, 55, 56^ and one tool tested only for reliability^57^.

### 3.3 Effectiveness in changing practitioner behaviour, knowledge, practice

Fourteen studies^34, 36, 37, 38, 39, 40, 41, 42, 45, 48, 50, 51, 58, 59^, described changes to practitioner behaviours, knowledge and/or practice in screening for child health behaviours (Table 3). Seven studies reported increased tool use and rates of screening^34, 36, 38, 39, 42, 48, 59^, three studies reported increased health-behaviour discussions/counselling^37, 40, 42^, and four studies reported improvements in health behaviour documentation^36, 37, 50, 58^. Further, three studies reported improved practitioner self-efficacy in addressing weight and health behaviours^40^, patient readiness to change^45^and addressing health behaviour goal setting^42^. Of the four studies that measured practitioner intention to use the tool in future, three reported moderate-high intention^41, 51, 59^.

### 3.4 Practitioner views on acceptability and feasibility of screening

Fourteen studies^35, 36, 37, 39, 40, 41, 42, 45, 48, 51, 52, 53, 58, 59^ described practitioner views on acceptability and/or feasibility of screening (Figure 2a; Supplementary Table 2). Common views positively impacting practitioner acceptability related to the value of screening^35, 37, 40, 41, 42, 45, 48, 51, 52, 53^ and features of the tool^40, 45, 51, 58, 59^ (Figure 2). Screening was commonly valued as being: useful or helpful in assessing health behaviours and facilitating health behaviour conversations with families; important; beneficial to families; and enhancing clinical sessions^41, 48, 52, 53^. Assorted screening tool features contributed to acceptability of screening, particularly simplicity and clarity^40, 45, 51, 58, 59^. Practitioners’ perceptions of feasibility were enhanced by the logistics of implementing screening, such as ease of use^35, 41^ and distribution^39^; ease to incorporate with clinic visits^48, 52^; and minimal impact on consultation time^37, 41, 48, 59^.

Conversely, negative practitioner perceptions on acceptability and feasibility related to the time required for screening, either undertaking screening or documenting outcomes in medical records^35, 36, 37, 45, 48, 51, 52^. Other factors limiting acceptability and feasibility related to caregiver difficulties completing screening or the wording of questions within the tools^36, 37, 51, 58^, disruption to workflow^45^, resourcing of IT infrastructure^58^, staffing capacity, skills and confidence^36, 37, 41, 45, 58^, or suitability of clinic type (i.e., not immunization clinic)^52^.

### 3.5 Caregiver views and acceptability on health behaviour screening tools

Eight studies^38, 41, 43, 45, 48, 49, 52, 53^ reported the views and acceptability of caregivers on health behaviour screening (Figure 2b, Supplementary Table 3). Caregivers were receptive to incorporating screening into the PHC setting^49^ valuing the opportunity to discuss health behaviours with their practitioner^45, 48^. Caregivers described being treated with care and feeling comfortable during consults with their practitioner^41, 45^, although some caregivers in one study reported a fear of being judged or appearing neglectful^49^. Caregivers across several studies were satisfied with the screening tool used and the resulting consultation^41, 45, 53^. Tools that were easy to use and took little time to read and complete were acceptable to caregivers^45, 49, 53^. Discussion of risk identification, goal setting and advice provided by practitioners following screening was well received, found to be useful and informative for caregivers^38, 41, 45, 49, 53^. Child acceptability was only discussed in one study: most caregivers and practitioners reported children were comfortable with the consultation, while some children experienced feelings of anxiety or demonstrated indifference^41^.

### 3.6 Training and resources needs

Eleven studies described practitioner-identified needs to support screening implementation^35, 38, 39, 40, 41, 45, 48, 50, 51, 52, 58^ (Table 5). These included: affordable provider/practitioner training and technical assistance^35, 39, 48, 50, 58^, practitioner resources to use alongside the screening tool such as referral pathways or behaviour change examples^38, 40, 41, 48, 51^, the integration of the screening tool into Electronic Medical Records^41, 45^, including reminders^48^, Dietitian support and/or follow up^48, 52^, patient (caregiver/child) educational resources^48^, and administrative support/capacity for implementation sustainability^40, 48^.

## 4. DISCUSSION

This systematic review identified and comprehensively described 14 unique child health behaviour screening tools used in PHC settings located across the United States, United Kingdom, and Canada. Screening tools measured health behaviours across the four domains of diet, physical activity, sedentary behaviour, and sleep, as well as related caregiving practices; however, only four screening tools included items across all four health behaviour domains. Screening tools were effective in changing practitioner self-reported behaviour, knowledge, self-efficacy, and provision of health behaviour education. The majority of included studies described practitioner or caregiver views on screening, indicating an overall high acceptability of health behaviour screening and feasibility within PHC. Training, resources, and integration into existing systems were identified as essential for implementation and screening success. However, given the heterogeneity in studies, a more comprehensive evaluation of the effectiveness, feasibility, and acceptability of screening tools within the PHC setting is called for.

Overall, this review identified a lack of brief, validated and reliable screening tools for use in the PHC setting that comprehensively measure child health behaviour globally. While four screening tools measured all four domains of diet, physical activity, sedentary behaviour and sleep, none were tested for validity or reliability. Similar to previous reviews examining health behaviour measurement tools^28, 29^, few tools focused on child sleep, indicating that sleep behaviours remain a comparatively novel area for early screening and intervention compared to diet and activity behaviours. This review demonstrated the effectiveness of screening tools in changing practitioner knowledge, attitudes and practice; but given that all studies used practitioner self-report measures, more robust evaluation of effectiveness are necessary to corroborate these findings.

Of the included studies, three-quarters reported on practitioner or caregiver acceptability and feasibility of screening, with most reporting positive indicators of acceptability and feasibility, such as finding screening tools valuable, easy to use and compatible with visits. Practitioners also indicated negative indicators of acceptability including time burden, limited staffing capacity, and incomplete and inconsistent completion of tools. Nonetheless, the depth of evaluation is limited. Heterogeneity in the evaluation designs, populations, data collection measures, reporting depth, and mixed findings of included studies, restricts our ability to draw firm conclusions on the acceptability and feasibility of screening from the current body of literature. For successful and sustained implementation of health behaviour screening in PHC settings, acceptability needs to be carefully evaluated from multiple perspectives including practitioners, support staff, practice managers, caregivers and children. Some studies included practice managers perspectives, and one study included caregiver-reported child perspectives, highlighting clear gaps. While screening was reported by practitioners and caregivers as valuable, feasibility may require further exploration as there were inconsistencies in practitioner views on the logistics of screening being easy to use versus time consuming to perform. Time burden is a particularly important consideration in PHC settings, due to existing time pressures and demand for existing priorities and responsibilities of PHC practitioners, including the treatment and management of disease and injury. As behaviour screening is proposed as a complementary practice to growth monitoring, time to conduct screening and undertake behaviour-directed conversations with caregivers needs to be appropriately resourced and funded. Given that studies often reported single aspects of acceptability or feasibility, or perspectives from only certain viewpoints, there is a need for future comprehensive assessment and co-design with key end-users to inform an acceptable and cost-effective implementation approach in PHC.

Challenges to implementing a change in routine practice include a lack of funding, resources, time and the need for administrative and managerial support^60^. Our review found a need to support PHC practices in these challenges, through providing adequate practitioner training and resources, integration into electronic medical records, administrative and dietitian support and patient education resources. Practitioners require adequate training to learn a new practice and feel confident and supported to implement the practice as part of their routine care. Literature suggests that it takes 17-20 years for the adoption of new interventions into routine practice^61^. This demonstrates that implementing a change in practice requires more than just screening tool dissemination, but a proactive and substantive collaboration with key stakeholders and the provision of adequate training and resources^62, 63^. This is supported by the findings of our review, which describes many practitioner-identified challenges to implementing a new practice of health behaviour screening. Practitioners identified training needs to support implementation and intervention success and highlighted the importance of integration of a screening tool into electronic medical records, staff roles and capacity and practitioner resources such as decision support charts, examples of specific behaviour change strategies and follow up consultations. This aligns with the findings of Krijger and colleagues^29^ who identified the importance and need for specific actions following screening that extend beyond counselling to address target behaviours, such as repeating screening after a certain time and referral to multidisciplinary team members. Qualitative literature also suggests engagement, open discussions and buy-in from PHC practitioners as vital to support adoption of new practices in PHC settings^64^. Successful implementation of health behaviour screening is achievable, but requires unique and adaptable end-user informed implementation strategies, tailored to the context and needs of the clinic, to support successful integration into PHC.

Key themes of Australian national public health policy include prioritising preventive health through screening and early intervention, indicating policy alignment for health behaviour screening as a potential early intervention and health promotion strategy^65, 66^. This review highlights several important avenues for future research that will be required to work towards policy directives regarding the implementation of screening and early intervention in PHC settings. While this review has identified several health behaviour screening tools that have been used in PHC, there is a lack of evidence regarding the validity and reliability of tools that assess all relevant health behaviour domains (i.e., nutrition, physical activity, sedentary behaviour and sleep). Prior to the implementation of health behaviour screening tools in PHC, the validity and reliability should be investigated to ensure the utility of these tools as screening instruments. Tools without sufficient validity and reliability may not be cost effective to implement^67^. The design of future research should be informed by a variety of end-users, including health practitioners, other PHC staff, caregivers and children. Collaborative engagement with end users would provide further insight into feasible and acceptable approaches to the implementation of health behaviour screening in PHC settings, as well as the support required to embed screening in routine care^68, 69^.

The results of this review should be considered in the context of strengths and limitations. The strengths include: (1) the review protocol being prospectively registered on PROSPERO with methodology according to PRISMA guidelines^30^, (2) the use of a comprehensive search strategy developed in collaboration with academic librarians across five databases, (3) contacting corresponding authors to retrieve screening tools not included in publications to enable complete assessment of screening tools. The primary limitation of this review is the exclusion of articles not published in English, grey literature, and unpublished theses, which may have limited inclusion of additional relevant literature or capturing of additional screening tools. Included studies also only came from the US, UK and Canada, limiting the generalisability to primary health care settings in other countries. The quality of included articles should also be recognised with most (17 of 22) included studies scoring 40% or lower using the MMAT critical appraisal tool, with Mixed Methods and Non-randomised studies being the most poorly reported. This highlights a lack of high-quality evidence within the limited body of literature regarding health behaviour screening in PHC.

## 5. CONCLUSION

Few screening tools exist to facilitate comprehensive screening of children’s health behaviours in PHC. Practitioners reported increased knowledge, self-efficacy, confidence and increased rates of documentation and health behaviour counselling, in addition to the barriers, enablers, training, and resource needs alongside screening tools. These findings provide new knowledge about the existence, implementation, acceptability, and feasibility of health behaviour screening tools, with mostly positive views. But the body of literature also demonstrates a need for more comprehensive evaluation and end-user informed implementation strategies to enable integration into PHC. This review highlights the potential of health behaviour screening as an acceptable and feasible strategy to comprehensively assess and provide early intervention for children’s health behaviours in PHC settings.

## Data Availability

The datasets used and/or analysed during the current study are available from the corresponding author on reasonable request.

## Acknowledgements

The authors would like to acknowledge the Flinders University research assistant team who supported this review: Alice Bradley, Samantha Pryde and Samantha Morgillo. The authors would also like to thank the Flinders University Research Librarian Mary Filsell who assisted with searching and the corresponding authors of included studies who responded to our request, Professor M. Diane McKee and Professor Michele Polacsek.

This work was supported by the Early Prevention of Obesity in Childhood, NMHRC Centre for Research Excellence (GNT1101675) and Translating Early Prevention of Obesity in Childhood, NMHRC Centre for Research Excellence (GNT2006999). DD and AM are supported by the Australian Government Research Training Program Scholarship. EH is supported by scholarships provided by the University of Sydney (UPA scholarship) and the Translating Early Prevention of Obesity in Childhood, NMHRC Centre for Research Excellence (GNT2006999).

## List of abbreviations

PHC: Primary Heath Care;
PRISMA: Preferred Reporting Items for Systematic Reviews and Meta-Analyses;
MMAT: Mixed Methods Appraisal Tool;
US: United States;
UK: United Kingdom;
N/R: Not reported;
SB: Sedentary Behaviour;
BMI: Body Mass Index;
PA: Physical Activity

## 6. DECLARATIONS

### Ethics approval and consent to participate

Not applicable. **Consent for publication** Not applicable.

### Competing interests

Competing interests: The authors have no competing interests.

### Authors contributions

RG, DZ, KD, EDW, BJ and LB conceived the project and provided study oversight. With the assistance of a research librarian, DZ developed the search strategy and DD conducted the search. DD, HC, RB, CR, DZ, KD and AM carried out article screening, DD conducted data extraction, and DD and EH completed critical appraisal. DD, HC, EH, BJ, LB and AM drafted the manuscript and all authors contributed to the interpretation of the results and critical review of the manuscript. All authors read and approved the final manuscript.

## Notes

### Competing Interest Statement

The authors have declared no competing interest.

