## Supplementary Tables for "Screening tools used in primary health care settings to identify health behaviours in children (birth – 16 years); A systematic review of their effectiveness, feasibility and acceptability"

**Supplementary File 1**

| Database | **Results** |
|---|---|
| MEDLINE | 1231 |
| PsycINFO | 593 |
| CINAHL | 2327 |
| Scopus | 1130 |
| Web of Science | 765 |

Example search in MEDLINE

Ovid®

Database(s): **Ovid MEDLINE(R) and Epub Ahead of Print, In-Process & Other Non-Indexed Citations, Daily and Versions(R)** 1946 to November 20, 2020

Search Strategy:

| # | Searches | Results |
|---|---|---|
| 1 | primary health care/ | 78919 |
| 2 | (primary care or primary medical care).tw. | 116915 |
| 3 | (primary health or primary healthcare).tw. | 30594 |
| 4 | general practice.tw. | 35628 |
| 5 | family practice/ | 65326 |
| 6 | (family practice or family medicine*).tw. | 17375 |
| 7 | (general practitioner* or gp* or general physician*).tw. | 221327 |
| 8 | (health* adj4 (provider* or personnel or worker* or profession*)).tw,kw. | 247111 |
| 9 | (family physician* or family doctor* or family practitioner*).tw. | 20772 |
| 10 | Health Personnel/ | 44900 |
| 11 | physicians, family/ | 16450 |
| 12 | or/1-11 | 680821 |
| 13 | community health services/ | 31873 |
| 14 | (communit* adj3 health).tw. | 51046 |
| 15 | 13 or 14 | 77702 |
| 16 | 12 or 15 | 736751 |
| 17 | exp Infant/ or exp Child/ or exp Child, Preschool/ or exp Pediatrics/ or exp Adolescent/ | 3615496 |
| 18 | (Child* or youth* or infant* or toddler* or "pre-school*" or infanc* or Adolescen* or teen* or Paediatric* or pediatric*).tw,kw. | 2102291 |
| 19 | 17 or 18 | 4152544 |
| 20 | exp Obesity/ or exp Pediatric Obesity/ | 216992 |
| 21 | (Obes* or over*weight or overweight or adipos* or "body fat*").tw,kw. | 417605 |
| 22 | 20 or 21 | 458178 |
| 23 | 19 and 22 | 104471 |
| 24 | Mass Screening/ | 104946 |
| 25 | "Surveys and Questionnaires"/ | 475182 |
| 26 | Qualitative Research/ | 58238 |
| 27 | Psychometrics/ | 76520 |
| 28 | "Diagnostic Techniques and Procedures"/ | 3368 |
| 29 | Decision Support Systems, Clinical/ or Decision Trees/ or Clinical Decision Rules/ or Clinical Decision-Making/ or Decision Making, Computer-Assisted/ or Decision Support Techniques/ | 50443 |
| 30 | (tool adj2 (screen* or test* or diagnos* or identi* or deci* or detect* or recog*)).kw,tw. | 75722 |
| 31 | (test adj2 (screen* or tool* or diagnos* or identi* or deci* or detect* or recog*)).kw,tw. | 72928 |
| 32 | (screen* adj4 (checklist or detect* or instrument* or index* or tool* or diagnos* or identi* or recog*)).kw,tw. | 120009 |
| 33 | (qualitative or "qualitative research").tw,kw. | 239390 |
| 34 | (checklist or detect* or instrument* or index or screen* or test* or diagnos* or identi* or deci* or detect* or recog*).kw,tw. | 10676173 |
| 35 | (earl* adj2 (detect* or ident* or screen* or diag* or recog*)).tw,kw. | 246764 |
| 36 | 24 or 25 or 26 or 27 or 28 or 29 or 30 or 31 or 32 or 33 or 35 | 1315629 |
| 37 | 36 and 23 and 16 | 1231 |

### Supplementary Table 1: MMAT Critical Appraisal

| Study | S1 | S2 | Qualitative Studies | | | | | Randomised Controlled Trials | | | | | Non-randomised Studies | | | | | Quantitative Descriptive Studies | | | | | Mixed Methods Studies | | | | | Final Score |
|---|---|---|---|---|---|---|---|---|---|---|---|---|---|---|---|---|---|---|---|---|---|---|---|---|---|---|---|---|
| *First author (Year)*<br>*Country*<br><br>*Study Design* | | | *1.1* | *1.2* | *1.3* | *1.4* | *1.5* | *2.1* | *2.2* | *2.3* | *2.4* | *2.5* | *3.1* | *3.2* | *3.3* | *3.4* | *3.5* | *4.1* | *4.2* | *4.3* | *4.4* | *4.5* | *5.1* | *5.2* | *5.3* | *5.4* | *5.5* | *Out of 100%* |
| Beno (2005)[35]<br>United States<br><br>Qualitative + Quantitative Descriptive | Y | Y | Y | UC | UC | N | UC | N/A | N/A | N/A | N/A | N/A | N/A | N/A | N/A | N/A | N/A | Y | N | UC | UC | UC | N/A | N/A | N/A | N/A | N/A | 20% |
| Hinchman (2005)[39]<br>United<br>States<br><br>Non-randomised study | Y | Y | N/A | N/A | N/A | N/A | N/A | N/A | N/A | N/A | N/A | N/A | UC | Y | Y | UC | UC | N/A | N/A | N/A | N/A | N/A | N/A | N/A | N/A | N/A | N/A | 40% |
| Dunlop (2007)[50]<br>United States<br><br>Quantitative Descriptive | Y | Y | N/A | N/A | N/A | N/A | N/A | N/A | N/A | N/A | N/A | N/A | N/A | N/A | N/A | N/A | N/A | Y | UC | Y | Y | Y | N/A | N/A | N/A | N/A | N/A | 80% |
| Woolford (2009)[51]<br>United States<br><br>Mixed Methods | Y | Y | Y | UC | UC | UC | UC | N/A | N/A | N/A | N/A | N/A | N/A | N/A | N/A | N/A | N/A | Y | N | Y | UC | Y | Y | N | N | N | UC | 20% |
| McKee (2010)[49]<br>United States<br><br>Qualitative | Y | Y | Y | Y | UC | Y | UC | N/A | N/A | N/A | N/A | N/A | N/A | N/A | N/A | N/A | N/A | N/A | N/A | N/A | N/A | N/A | N/A | N/A | N/A | N/A | N/A | 60% |
| Watson-Jarvis (2011a)[52]<br>Canada<br><br>Quantitative Descriptive | Y | Y | N/A | N/A | N/A | N/A | N/A | N/A | N/A | N/A | N/A | N/A | N/A | N/A | N/A | N/A | N/A | Y | UC | UC | N | UC | N/A | N/A | N/A | N/A | N/A | 20% |
| Watson-Jarvis (2011b)[53]<br>Canada<br><br>Quantitative Descriptive | Y | Y | N/A | N/A | N/A | N/A | N/A | N/A | N/A | N/A | N/A | N/A | N/A | N/A | N/A | N/A | N/A | Y | N | Y | N | Y | N/A | N/A | N/A | N/A | N/A | 60% |
| Andrade (2020)[48]<br>Canada<br><br>Mixed Methods | Y | Y | Y | UC | UC | Y | UC | N/A | N/A | N/A | N/A | N/A | N/A | N/A | N/A | N/A | N/A | Y | N | UC | UC | Y | Y | Y | UC | UC | UC | 40% |
| Christison (2014)[45]<br>United States<br><br>Non-randomised Study | Y | Y | N/A | N/A | N/A | N/A | N/A | N/A | N/A | N/A | N/A | N/A | N | N | UC | UC | UC | N/A | N/A | N/A | N/A | N/A | N/A | N/A | N/A | N/A | N/A | 20% |
| Herbenick (2018)[46]<br>United States<br><br>Quantitative Descriptive | Y | Y | N/A | N/A | N/A | N/A | N/A | N/A | N/A | N/A | N/A | N/A | N/A | N/A | N/A | N/A | N/A | Y | UC | UC | N | N | N/A | N/A | N/A | N/A | N/A | 20% |
| Bailey-Davis (2019)[34]<br>United States<br><br>Non-randomised Study | Y | Y | N/A | N/A | N/A | N/A | N/A | N/A | N/A | N/A | N/A | N/A | Y | UC | Y | N | UC | N/A | N/A | N/A | N/A | N/A | N/A | N/A | N/A | N/A | N/A | 40% |
| Gance-Cleveland (2014)[58]<br>United States<br><br>Non-randomised Study | Y | Y | N/A | N/A | N/A | N/A | N/A | N/A | N/A | N/A | N/A | N/A | UC | Y | UC | UC | UC | N/A | N/A | N/A | N/A | N/A | N/A | N/A | N/A | N/A | N/A | 20% |
| Park (2015)[41]<br>United Kingdom | Y | Y | Y | Y | UC | UC | UC | N/A | N/A | N/A | N/A | N/A | N | UC | UC | Y | UC | N/A | N/A | N/A | N/A | N/A | N/A | N/A | N/A | N/A | N/A | 20% |

| | | | | | | | | | | | | | | | | | | | | | | | | | | | | |
|---|---|---|---|---|---|---|---|---|---|---|---|---|---|---|---|---|---|---|---|---|---|---|---|---|---|---|---|---|
| Mixed Methods | | | | | | | | | | | | | | | | | | | | | | | | | | | | |
| Sharpe (2016)[43]<br>United States<br><br>Non-randomised Study | Y | Y | N/A | N/A | N/A | N/A | N/A | N/A | N/A | N/A | N/A | N/A | UC | UC | N | UC | UC | N/A | N/A | N/A | N/A | N/A | N/A | N/A | N/A | N/A | N/A | 20% |
| Polacsek (2009)[42]<br>United States<br><br>Non-randomised study | Y | Y | N/A | N/A | N/A | N/A | N/A | N/A | N/A | N/A | N/A | N/A | UC | UC | Y | N | UC | N/A | N/A | N/A | N/A | N/A | N/A | N/A | N/A | N/A | N/A | 20% |
| Gibson (2016)[38]<br>United States<br><br>Quantitative Descriptive | Y | Y | N/A | N/A | N/A | N/A | N/A | N/A | N/A | N/A | N/A | N/A | N/A | N/A | N/A | N/A | N/A | Y | Y | UC | UC | Y | N/A | N/A | N/A | N/A | N/A | 60% |
| Camp (2017)[37]<br>United States<br><br>Mixed Methods | Y | Y | Y | UC | UC | N | UC | N/A | N/A | N/A | N/A | N/A | Y | UC | Y | N | UC | N/A | N/A | N/A | N/A | N/A | N | N | UC | UC | UC | 20% |
| Camp (2020)[36]<br>United States<br><br>Mixed Methods | Y | Y | Y | UC | UC | UC | UC | N/A | N/A | N/A | N/A | N/A | N/A | N/A | N/A | N/A | N/A | Y | UC | UC | Y | Y | N | N | UC | UC | UC | 20% |
| Karacabeyli (2020)[40]<br>Canada<br><br>Mixed Methods | Y | Y | Y | Y | UC | Y | UC | N/A | N/A | N/A | N/A | N/A | UC | UC | UC | UC | UC | N/A | N/A | N/A | N/A | N/A | Y | U/C | U/C | N | N | 20% |
| Savage (2018)[47]<br>United States<br><br>Randomised Controlled Trial Protocol | Y | Y | N/A | N/A | N/A | N/A | N/A | UC | UC | UC | UC | UC | N/A | N/A | N/A | N/A | N/A | N/A | N/A | N/A | N/A | N/A | N/A | N/A | N/A | N/A | N/A | 20% |
| Shook (2018)[44]<br>United States<br><br>Quantitative Descriptive | Y | Y | N/A | N/A | N/A | N/A | N/A | N/A | N/A | N/A | N/A | N/A | N/A | N/A | N/A | N/A | N/A | Y | N | Y | Y | Y | N/A | N/A | N/A | N/A | N/A | 80% |
| Williams (2020)[59]<br>United States<br><br>Mixed Methods | Y | Y | Y | UC | UC | Y | UC | N/A | N/A | N/A | N/A | N/A | N/A | N/A | N/A | N/A | N/A | Y | N | UC | Y | Y | Y | N | UC | UC | UC | 20% |

Abbreviations: Y: Yes, N: No, N/A: Not applicable, UC: Unclear
MMAT, Mixed Methods Assessment Tool[33], MMAT scored out of 100%, 20% per question, higher % score indicating higher quality study

#### Supplementary Table 2: Practitioner views on acceptability and feasibility of screening

| | |
|---|---|
| **Value of screening** | • Useful and effective for patient care[35]<br>• Useful or very useful with patients[42]<br>• Enabled assessment and benefited families[52]<br>• Valued the screen and felt it enhanced the visit[48]<br>• Screening is important[52, 53]<br>• Somewhat or very helpful in assessing and communicating weight-related risk factors[51]<br>• Helpful in providing weight management recommendations[51]<br>• Facilitated healthy eating/weight conversations[48]<br>• Tool is useful and practitioners liked the tool[45]<br>• Tool was useful or somewhat useful, would recommend the tool to other health professionals, and improved their ability to care for the child[41]<br>• Improved dietary and activity assessment and facilitated engagement with caregivers about their child's health habits[37]<br>• Messaging of resources facilitated practice change and empowered practitioners to be proactive with health promotion[40] |
| **Features of tool** | • Tool was attractive and helpful for caregivers[51]<br>• Tool is accurate[45]<br>• Simplicity and clarity of tool message[40]<br>• Interview (i.e., screening) and printed summary functioned as good discussion aids[58]<br>• Tool standardizes, facilitates and streamlines healthy lifestyle conservations with families[59] |
| **Logistics** | • Helpful and easy to use[35]<br>• Easy to distribute to patients[39]<br>• Incorporating screening into clinic was easy[52]<br>• Screening was compatible with visits[48]<br>• Tool was easy/ straightforward to use, tool saved them time[41]<br>• Caregiverss completed screen in the waiting room pre-consultation[48]<br>• No increase in time needed to use tool[37]<br>• Able to use screening tool consistently[36]<br>• Tool reduced or did not significantly add to practitioner cognitive workload[59] |
| **Time** | • Time consuming[35]<br>• Time was the most frequently mentioned barrier[51]<br>• Common challenge was time[52]<br>• Additional time required[48]<br>• Time to use and increased appointment duration[45] |

| | |
|---|---|
| | • Electronic documentation of tool into EMR was time consuming[36, 37] |
| **Ease of caregiver/child competition** | • Tool wording occasionally confusing for patients[51]<br>• Some caregivers had difficulty completing screen[37]<br>• Not always completed or completed fully[36]<br>• Younger students (i.e., participants) needed extra help completing the interview (i.e., screening)[58] |
| **Change in practice required** | • Caused disruption to workflow[45]<br>• Lack of existing IT infrastructure, limited clinical IT support and provider IT skills/discomfort with IT[58]<br>• Limited staffing and resistance to change[58]<br>• Tool required some practice, feeling uncomfortable discussing child weight-related health risk with caregivers[41]<br>• Inconsistency with handout distribution by nursing staff[36, 37]<br>• Immunization clinic was not a convenient location to administer tool – caregiver engagement and time[52] |

**Supplementary Table 3: Caregiver views on health behaviour screening tools**

| Study | Caregiver views |
|---|---|
| *First author (Year)*<br>*Country*<br><br>*Tool Name* | *Caregiver acceptability* |
| McKee (2010)[49]<br>United States<br><br>The Family Lifestyle Assessment of Initial Risk (FLAIR) screening form | • All families agreed that assessing health behaviors should be part of well-child visits.<br>• Tool was easy to complete and something that should continue.<br>• Fear of being judged or appearing neglectful.<br>• Importance of doctor's involvement in screening.<br>• Positive overall impression of the goal setting and lifestyle counseling.<br>• Appreciated variety of accompanying resources including pamphlets, recipes and websites. |
| Watson-Jarvis (2011a)[52]<br>Canada<br><br>Nutrition Screening Tool for Every Preschooler (NutriSTEP) | • 'Easy' or 'very easy' to complete (99%)<br>• 'Moderately' or 'very helpful' for identifying areas of nutrition concern (77%)<br>• Not very helpful (18%)<br>• 'Moderately' or 'very interested' in completing screen in health centre (84%) or practitioners office (81%)<br>• Clerks identified caregiver concern about the amount of reading required |
| Watson-Jarvis (2011b)[53]<br>Canada<br><br>Nutrition Screening Tool for Every Preschooler (NutriSTEP) | 63% of caregivers were satisfied with the service and 38% had a neutral opinion |
| Andrade (2020)[48]<br>Canada<br><br>Nutrition Screening Tool for Every Preschooler (NutriSTEP) | Practitioners reported caregivers appreciated the opportunity to discuss nutrition related issues with practitioners at their scheduled appointments, regardless of their child's nutritional risk score. |
| Christison (2014)[45]<br>United States<br><br>The Family Nutrition and Physical Activity (FNPA) risk assessment tool | Satisfaction survey (5-point Likert scale)<br>• Caregiver satisfaction with the tool was high<br>• Tool was easy to read, easy to fill out and little time to complete<br>• Discussion with provider was helpful, important, made caregivers feel comfortable, right amount of time and felt practitioner listened<br>• Lower scores for motivating family and child change |
| Park (2015)[41]<br>United Kingdom<br><br>Computer-Assisted Treatment of CHildhood overweight (CATCH) | • All caregivers (n=14) reported that they and their child felt comfortable with the consultation and being asked about their child's lifestyle and medical history<br>• Caregivers were satisfied (n=12) or 'somewhat satisfied (n=2) with the tool-aided consultation<br>• One caregiver was 'slightly uncomfortable' when asked about whether their child had been teased/bullied.<br>• Caregivers found it 'useful' (n=11) or 'somewhat useful' (n=3) to receive personalised feedback<br>• All caregivers agreed that they were treated with care and concern, that their child's care was well organized and that they had confidence and trust in their practitioner. |

| | |
|---|---|
| | • Consults described as positive, informative, nonjudgmental, and nonintrusive.<br>• Caregivers found the tool's outputs useful.<br>• Two caregivers described consultation causing some anxiety in their children.<br>• Caregivers found the lifestyle advice informative and instructive, particularly specific advice on diet as being useful.<br>• Follow-up appointments for monitoring, guidance and practical support would be beneficial (n=5) |
| Sharpe (2016)[43]<br>United States<br><br>Starting the Conversations (STC) 4-12 tool | • Discussion helped motivate entire family to make healthier changes<br>• One behaviour change goal empowered families to set achievable goals and avoid feeling overwhelmed |
| Gibson (2016)[38]<br>United States<br><br>Healthy Habits Questionnaire | Tool heightened caregiver awareness of the lifestyle habits of the family and motivated the caregiver to make changes in their diet and physical activity |
